# Implications of the 2026 Cardiovascular-Kidney-Metabolic Syndrome Guideline for GLP-1 Receptor Agonist and SGLT2 Inhibitor Eligibility Among US Adults

**DOI:** 10.64898/2026.10.22.26366274

**Authors:** Douglas R. Corsi, Calvin Wang, Yu Mi Kang, André Zimerman, Andrew O. Agbaje, Filipe A. Moura

## Abstract

**Background:** The 2026 cardiovascular-kidney-metabolic (CKM) syndrome guideline broadens recommendations for glucagon-like peptide-1 receptor agonists (GLP-1RA) and sodium-glucose cotransporter 2 inhibitors (SGLT2i), but the increment over pre-2026 standards is unknown. We estimated the number and clinical profile of US adults newly eligible under this guideline.

**Methods:** Cross-sectional analysis of adults aged 30-79 years in the 2011-2018 National Health and Nutrition Examination Survey. Adults were newly eligible if they met 2026 CKM guideline indications but not pre-2026 American Diabetes Association, American College of Cardiology/American Heart Association heart failure, Kidney Disease: Improving Global Outcomes, or obesity guideline indications. Sensitivity analyses used 2 US Food and Drug Administration (FDA) label standards.

**Results:** Among 175.17 million US adults, 111.84 million (63.8%) were already eligible for either class. The CKM guideline added 7.48 million (95% CI: 6.48 to 8.47), a 6.7% increase, all for GLP-1RA and none for SGLT2i: 6.49 million with overweight (body mass index 27.0-29.9 kg/m2, ethnicity adjusted) and no other cardiometabolic disease, and 0.99 million with type 2 diabetes and 10-year PREVENT cardiovascular risk ≥7.5%. Newly eligible adults (86.8% CKM stage 1) were younger (46.2 vs 52.9 years), leaner (27.8 vs 33.0 kg/m2), and at lower 10- and 30-year PREVENT risk (3.6% vs 8.1%; 11.3% vs 19.6%; all P < 0.001). Against FDA labeling, 14.15-25.17 million were newly eligible (7.66-18.20 million for SGLT2i). Assuming a 20% relative risk reduction, the projected number needed to treat per cardiovascular event prevented was 243 over 10 years and 56 over 30 years (ages 30-59) in adults with overweight, versus 61 and 26 in already eligible adults.

**Conclusions:** Relative to pre-2026 society guidelines, the 2026 CKM guideline newly confers a GLP-1RA indication on 7.48 million US adults, predominantly at lower cardiovascular risk. Whether treating these adults improves outcomes in a cost-efficient manner remains to be established.

**What Is Known:**

- The 2026 AHA/ACC/ADA/ASN cardiovascular-kidney-metabolic (CKM) syndrome guideline recommends GLP-1 receptor agonists and SGLT2 inhibitors according to CKM stage, comorbidity, and PREVENT-predicted risk, including a GLP-1 receptor agonist for adults with overweight at CKM stage 1.
- National estimates of eligibility for these agents predate the guideline and have largely used FDA labeling, rather than professional society guidance, as the reference standard.

**What the Study Adds:**

- Relative to pre-2026 society guidance, the guideline newly identifies 7.48 million US adults aged 30 to 79 years as treatment candidates, all for a GLP-1 receptor agonist and none for an SGLT2 inhibitor; most have overweight without other cardiometabolic disease.
- Newly eligible adults are younger and at lower predicted cardiovascular risk than those already eligible; the projected number needed to treat is 243 over 10 years and 56 over 30 years, so expected benefit rests on a lifetime horizon that outcome trials and cost-effectiveness analyses have not yet tested.

## INTRODUCTION

Cardiovascular-kidney-metabolic (CKM) syndrome links adiposity, metabolic risk factors, chronic kidney disease (CKD), and cardiovascular disease across a staging spectrum from no CKM risk factors (stage 0) to established clinical cardiovascular disease (stage 4).^1^ Roughly 9 in 10 US adults are at stage 1 or higher.^2^ The 2026 CKM guideline recommends glucagon-like peptide-1 receptor agonists (GLP-1RA) and sodium-glucose cotransporter 2 inhibitors (SGLT2i) according to CKM stage and comorbidity.

The 2026 CKM guideline broadens the populations for whom these agents are recommended, and we examined four cases of indication expansion.^3^ First, it recommends a GLP-1RA for adults with overweight (body mass index [BMI] 27.0 to 29.9 kg/m2, ethnicity-adjusted) at CKM stage 1, in the absence of any weight-related comorbidity. Second, it recommends adding a GLP-1RA in type 2 diabetes with CKD and a urine albumin-to-creatinine ratio (UACR) of 100 mg/g or greater. Third, the guideline anchors initiation to predicted absolute risk rather than to prevalent disease: adults with type 2 diabetes at CKM stage 2 or 3 whose 10-year PREVENT total-cardiovascular disease (CVD) risk is 7.5% or greater are recommended a GLP-1RA, an SGLT2i, or both, even without established cardiovascular or kidney disease. Fourth, it recommends an SGLT2i for adults without diabetes who have CKD with a UACR of 30 to 199 mg/g and an estimated glomerular filtration rate (eGFR) of 20 mL/min/1.73 m2 or greater, below the albuminuria thresholds that defined entry to the pivotal kidney outcome trials.^4,5^

Whether any of these recommendations identifies a new treatment candidate depends on the guideline used as the comparator, and the available reference standards diverge. US Food and Drug Administration (FDA) labeling, under which type 2 diabetes alone confers an indication for both classes, is broader than professional society guidance in some respects and narrower in others. Previous studies have used the National Health and Nutrition Examination Survey (NHANES) to derive national eligibility estimates for cardiometabolic therapies after new guideline releases.^6–9^ However, existing national estimates of eligibility for GLP-1RA and SGLT2i predate the 2026 CKM treatment recommendations and have largely used FDA labeling rather than contemporary professional society guidance as the comparator.^9^ Consequently, the extent to which the 2026 CKM guideline identifies additional treatment candidates, and which clinical populations account for that expansion, remains unknown.

Using NHANES 2011 to 2018, we therefore estimated the number and clinical characteristics of US adults newly eligible for a GLP-1RA, SGLT2i, or both under the 2026 CKM guideline, taking contemporary professional society guidance as the primary reference standard and FDA labeling as a sensitivity analysis.

## METHODS

### Data Source and Study Population

We conducted a serial cross-sectional analysis of the NHANES, a stratified, multistage probability sample of the civilian, noninstitutionalized US population conducted by the National Center for Health Statistics.^10^ We pooled four 2-year cycles from 2011 to 2018 and applied 8-year mobile examination center (MEC) weights, accounting for the complex survey design. Data were analyzed from June to August 2026. Complete survey-design specifications, including the weight, stratum, and primary sampling unit variables used, are provided in the Supplemental Methods. The NHANES protocol was approved by the National Center for Health Statistics Research Ethics Review Board, and all participants provided written informed consent. Because this secondary analysis used deidentified, publicly available data, it was exempt from further institutional review board review under the Common Rule (45 CFR §46). We followed the Strengthening the Reporting of Observational Studies in Epidemiology (STROBE) reporting guideline.^11^

We first restricted the pooled release to nonpregnant adults aged 20 years or older with a valid MEC examination weight, and then to those aged 30 to 79 years, the age range targeted by the AHA PREVENT equations and by the 2026 CKM guideline’s primary-prevention risk recommendations. Participants were required to have the measurements necessary for CKM staging (anthropometry, blood pressure, glycemia, lipids, serum creatinine, and urinary albumin-to-creatinine ratio). Derivation of the analytic sample from the pooled NHANES release is shown in Figure S1 (study flow). The final analytic sample comprised 16,341 participants representing 175.17 million US adults. The staging algorithm reproduced published national estimates of CKM stage prevalence (Table S1).^2^

### CKM Syndrome Staging

We assigned each participant a CKM stage (0–4) following the 2026 CKM guideline.^3^ Detailed stage definitions, adapted to the data available in NHANES, are given in Table S2, Part E.^12,13^

### Assessment of Clinical Variables and Risk Estimation

Type 2 diabetes was identified by self-reported diagnosis, hemoglobin A1c ≥6.5%, fasting plasma glucose ≥126 mg/dL, or use of glucose-lowering medication. Estimated glomerular filtration rate (eGFR) was calculated from serum creatinine using the 2021 Chronic Kidney Disease Epidemiology Collaboration (CKD-EPI) creatinine equation,^14^ and the urinary albumin-to-creatinine ratio (UACR) was taken from a single spot urine specimen. CKD was defined by Kidney Disease: Improving Global Outcomes (KDIGO) criteria as an eGFR <60 mL/min/1.73 m² or a UACR ≥30 mg/g.^15^ Established atherosclerotic cardiovascular disease (ASCVD) was defined from self-report of coronary heart disease, myocardial infarction, stroke, or angina. Heart failure was defined by self-report of heart failure of any type. Because ejection fraction is not measured in NHANES, recommendations specific to heart failure with preserved ejection fraction were not operationalized. Ten-year risks of total cardiovascular disease, ASCVD, and heart failure were estimated using the AHA Predicting Risk of Cardiovascular Disease EVENTs (PREVENT)^16^ equations; corresponding 30-year risks were additionally estimated for adults aged 30 to 59 years, the age range for which the 30-year equations are defined.

### 2026 Guideline Indications (Analytic Cohorts)

We operationalized four 2026 CKM guideline recommendations, each defining a cohort of adults for whom the CKM guideline recommends a GLP-1RA and/or SGLT2i. Exact recommendation text, class of recommendation, and level of evidence are given in Table S3. Adults with overweight and no other cardiometabolic disease (GLP-1RA) are identified by the indication for GLP-1RA in overweight, defined as a BMI of 27.0 to 29.9 kg/m², or 25.0 to 27.4 kg/m² for adults of self-reported Asian ancestry;^12^ this ethnicity-adjusted band is the one referred to as “overweight” throughout. Adults with type 2 diabetes and albuminuria (GLP-1RA add-on) are identified by the indication for GLP-1RA in type 2 diabetes with CKD, UACR ≥100 mg/g, at CKM stage 2–3. Adults with type 2 diabetes at the risk threshold (GLP-1RA and SGLT2i) are identified by the recommendation for GLP-1RA or SGLT2i in type 2 diabetes at CKM stage 2–3 with 10-year PREVENT total-CVD risk ≥7.5%, without established ASCVD, heart failure, or CKD. Adults with CKD without diabetes (SGLT2i) are identified by the indication for an SGLT2i in CKD without diabetes but with UACR 30–199 mg/g and eGFR ≥20 mL/min/1.73 m² (KDIGO categories G1–G4, A2).

### Comparator Reference Standards

For each cohort we defined “newly eligible” as meeting the CKM guideline indication while not meeting the comparator indication for the same drug class. Throughout, an adult is described as eligible when they meet the criteria of the standard under discussion, whether that standard is a guideline recommendation or an FDA label. All comparator predicates were operationalized in NHANES with explicit cutoffs (Table S3); complete operational definitions are provided in Table S2. FDA-approved indications were compiled for each product in both classes from the manufacturer prescribing information accessed via accessdata.fda.gov (Table S2, Part D):

#### Main analysis: Comparison against composite society guidelines

An adult was considered already eligible if indicated by any major pre-2026 professional society guideline: type 2 diabetes with ASCVD or broad CKD, or American Diabetes Association (ADA) high-risk type 2 diabetes^17^; heart failure of any type (2022 AHA/ACC/HFSA)^18^; broad CKD by KDIGO (eGFR <60 mL/min/1.73 m² or UACR ≥30 mg/g)^15^; or obesity per American Association of Clinical Endocrinology^19^/American Gastroenterological Association^20^ weight-management criteria (BMI ≥30 kg/m², or 27.0 to 29.9 kg/m² with a weight-related comorbidity). ADA high-risk type 2 diabetes was defined as age ≥55 years and ≥2 of obesity (BMI ≥30 kg/m²), hypertension, current smoking, dyslipidemia, or albuminuria (UACR ≥30 mg/g).

#### Comparison against Full FDA label (including the glycemic indication, sensitivity analysis 1)

Type 2 diabetes alone confers an indication for both classes, in addition to heart failure and trial/label-threshold CKD (both classes’ disease-based indications).

#### Comparison against FDA label restricted to compelling cardiorenal disease (Sensitivity analysis 2)

We used the term compelling cardiorenal disease to denote comorbidities with well-established indications for GLP-1RA or SGLT2i therapy. These included established ASCVD, heart failure, or trial/label-threshold CKD. Type 2 diabetes confers an indication under this standard only in the presence of one of these three conditions. Trial/label-threshold CKD was defined as eGFR ≥20 and UACR ≥200 mg/g or eGFR <45 mL/min/1.73 m², the union of the pivotal CKD trials that enrolled adults without diabetes that anchor the SGLT2i label (EMPA-KIDNEY, DAPA-CKD).^4,5^ This comparator is not intended to describe FDA labeling as written, under which type 2 diabetes alone permits both classes, but to isolate the CKM guideline’s cardiorenal rationale from its glycemic one.

The three comparators are not nested overall. For GLP-1RA, FDA labeling was broadest because type 2 diabetes alone qualifies as an indication, whereas for SGLT2i, society guidance was broadest because it included KDIGO-defined CKD. Accordingly, cross-comparator differences were described rather than formally tested. We also assessed the union of all three comparators, considering participants already eligible if any pre-2026 society guideline or FDA label covered that drug class.

### Statistical Analysis

Estimates incorporated the NHANES survey design using Taylor-series linearization (Supplemental Methods). We report weighted counts, weighted percentages, and unweighted sample sizes, each with 95% confidence intervals. Population counts were derived from weighted proportions and NHANES population totals. Newly eligible and already-eligible adults were compared using Rao-Scott χ² tests for categorical variables and survey-weighted Kruskal-Wallis tests for continuous variables. Continuous variables are reported as weighted means (95% CIs), except UACR, reported as median (interquartile range). Cross-comparator differences were not formally tested. For each drug class we report the relative increase, defined as the number of newly eligible adults as a percentage of the already-eligible population under the same standard. Eligibility was also tabulated by drug class and CKM stage. Projected numbers needed to treat over 10 and 30 years were calculated as 1/(mean predicted PREVENT risk × 0.20), using the guideline’s assumed 20% relative risk reduction;^21^ confidence intervals were derived from the confidence limits of mean risk. Total CVD was the primary endpoint; ASCVD was also reported to match trial-derived effect estimates. Projections were restricted to participants with computable PREVENT risk, with 30-year estimates limited to ages 30 to 59 years. These projections assume full uptake and adherence and extrapolate trial effects to populations not enrolled in those trials. Analyses used R version 4.5.2; two-sided P < 0.05 defined statistical significance.

### Data Availability

NHANES data are publicly available from the National Center for Health Statistics. The analysis code is available from the corresponding author on reasonable request. Dr Corsi had full access to all the data in the study and takes responsibility for its integrity and the data analysis.

## RESULTS

The weighted study sample included 16,341 NHANES participants representative of 175.17 million US adults aged 30 to 79 years (weighted mean age, 51.5 years; 51.8% female; 5.6% non-Hispanic Asian, 11.1% non-Hispanic Black, 14.2% Hispanic, 65.8% non-Hispanic White, 3.4% other or multiracial). Mean BMI was 29.8 kg/m², and the prevalences of type 2 diabetes, hypertension, dyslipidemia, and metabolic syndrome were 16.6%, 58.3%, 42.2%, and 21.8%, respectively; 12.8% met KDIGO criteria for chronic kidney disease, 8.3% reported established ASCVD, and 2.5% reported heart failure. CKM stage distribution was 8.1%, 23.2%, 55.0%, 4.6%, and 9.1% for stages 0 through 4 (Table S4). Before the 2026 CKM guideline was applied, 111.84 million US adults (63.8%) were already eligible for at least 1 of the 2 drug classes under the composite society standard (Table 1): 105.10 million (60.2%) for a GLP-1RA and 44.68 million (25.5%) for an SGLT2i. The corresponding FDA counts appear in Tables S5 and S6.

**Table 1.**
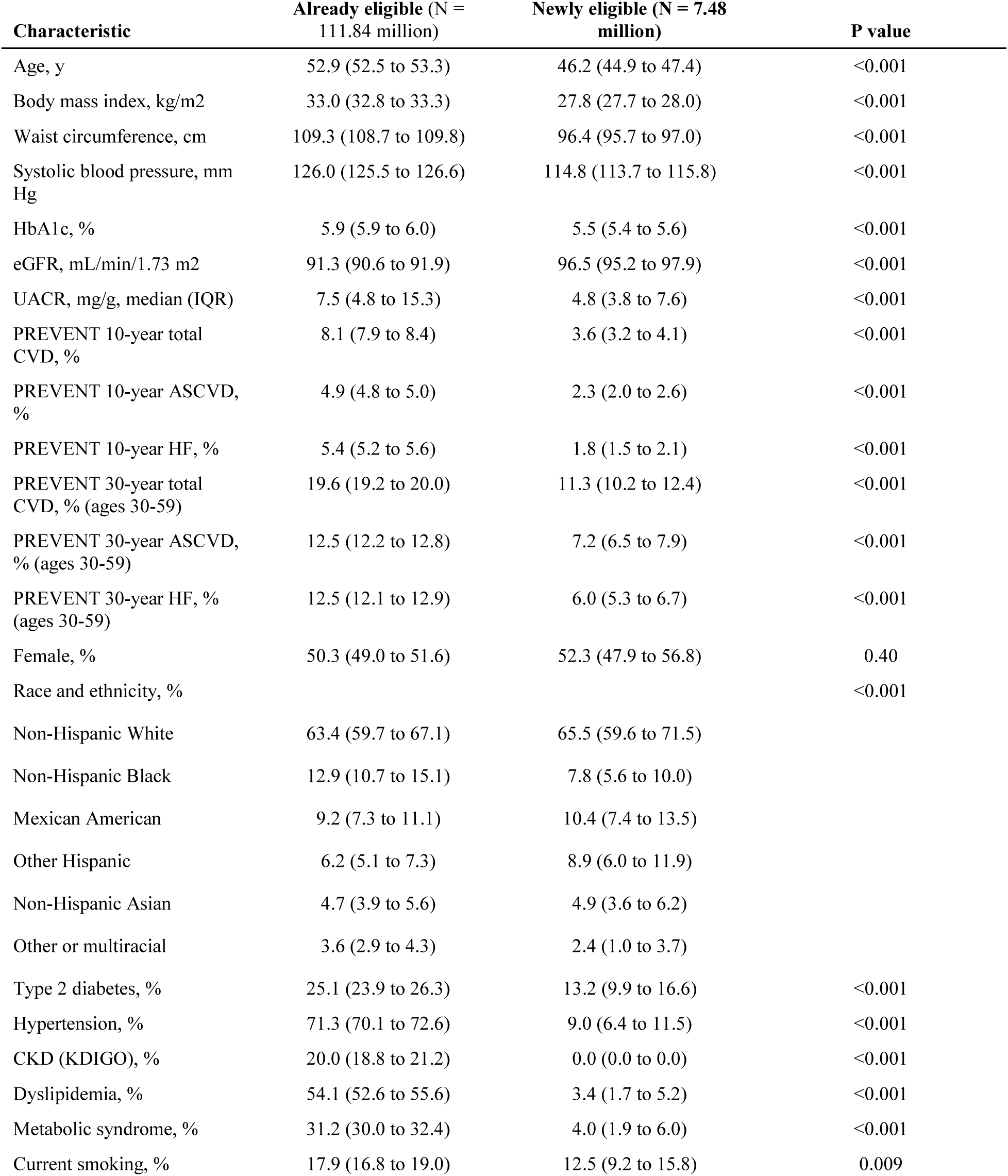
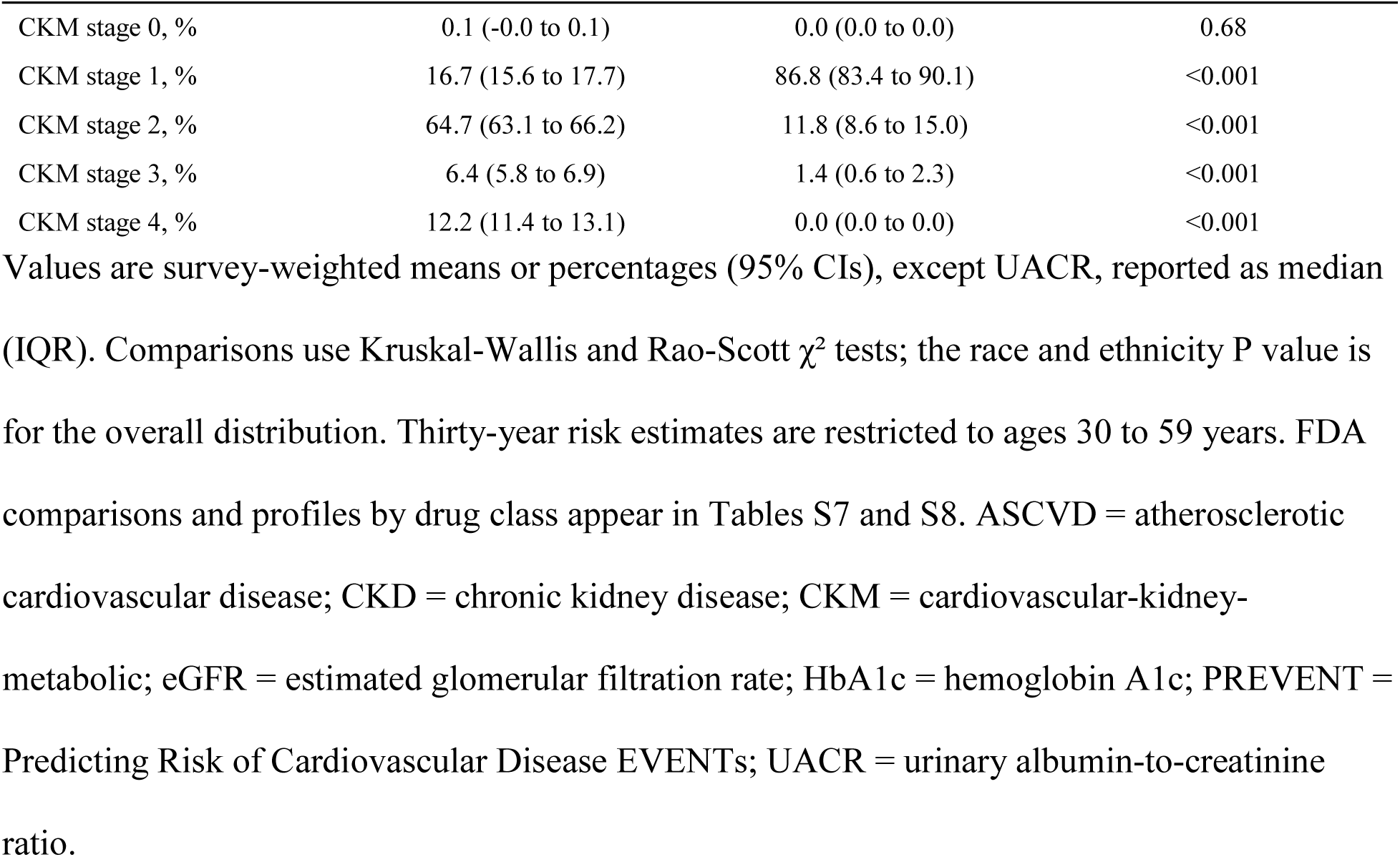
Clinical profile of adults newly eligible under the 2026 CKM guideline vs society guidance. Values are survey-weighted means or percentages (95% CIs), except UACR, reported as median (IQR). Comparisons use Kruskal-Wallis and Rao-Scott χ² tests; the race and ethnicity P value is for the overall distribution. Thirty-year risk estimates are restricted to ages 30 to 59 years. FDA comparisons and profiles by drug class appear in Tables S7 and S8. ASCVD = atherosclerotic cardiovascular disease; CKD = chronic kidney disease; CKM = cardiovascular-kidney-metabolic; eGFR = estimated glomerular filtration rate; HbA1c = hemoglobin A1c; PREVENT = Predicting Risk of Cardiovascular Disease EVENTs; UACR = urinary albumin-to-creatinine ratio.

### Newly Eligible Adults Under the 2026 CKM Guideline: Main Analysis (Composite Society Comparator)

Compared with the composite of major pre-2026 professional society guidelines, an estimated 7.48 million US adults aged 30 to 79 years (4.28%; 95% CI: 3.80 to 4.76; unweighted n = 620) were newly eligible for a GLP-1RA or an SGLT2i under the 2026 CKM guideline, a 6.7% relative increase over the 111.84 million already eligible (Figure 1; Table 2). All were GLP-1RA candidates, a 7.1% increase in the GLP-1RA-eligible population: 6.49 million adults (95% CI: 5.51 to 7.47) with overweight (BMI 27.0 to 29.9 kg/m²) and no other cardiometabolic disease, all at CKM stage 1, plus 0.99 million adults (95% CI: 0.75 to 1.23) with type 2 diabetes who met the CKM guideline’s PREVENT-based risk threshold but did not qualify as ADA high-risk. No adults were newly eligible for SGLT2i therapy under the society comparator because the CKD cohort met the KDIGO albuminuria criterion (UACR ≥30 mg/g).

**Figure 1.**
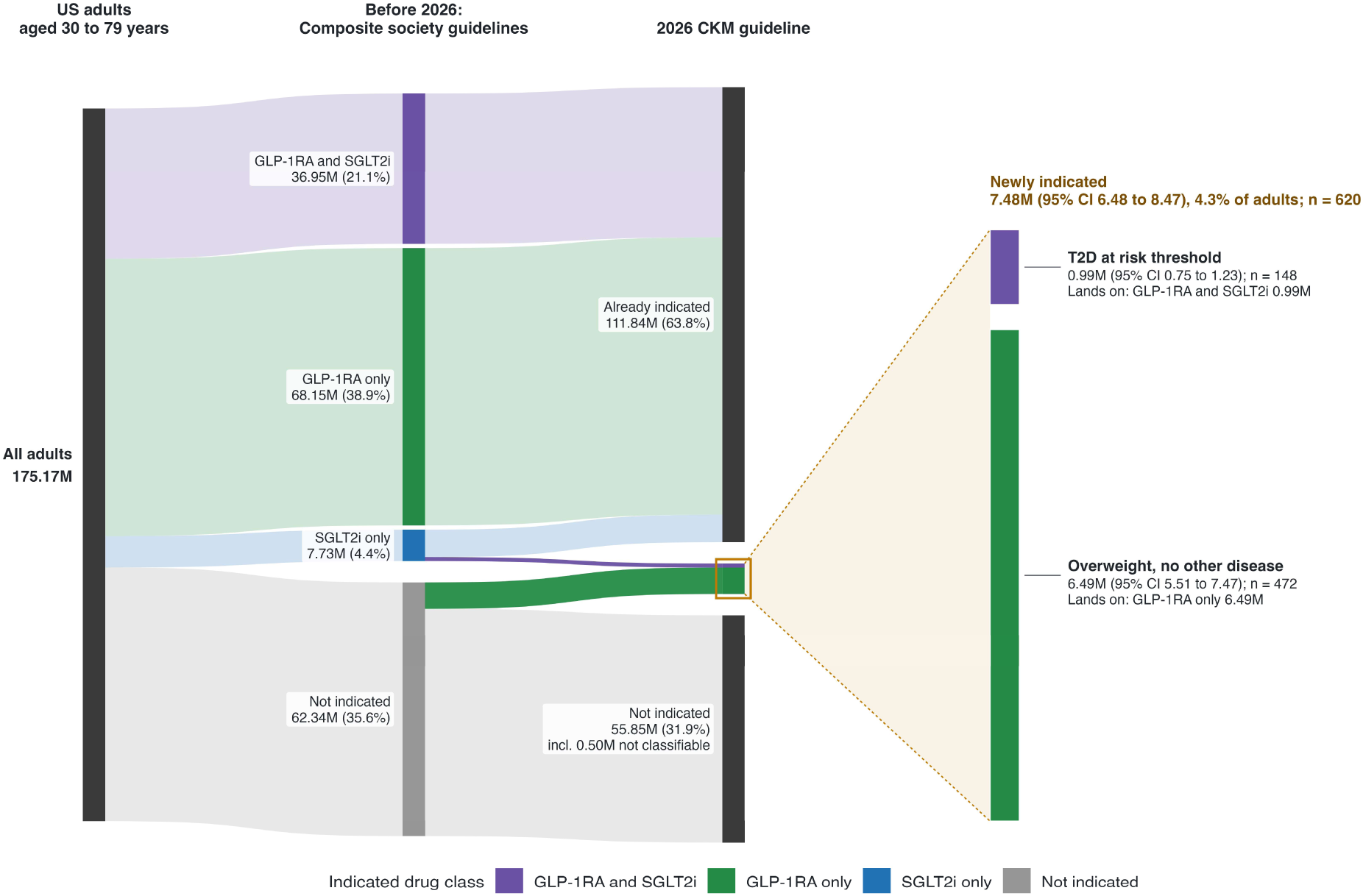
Reclassification from pre-2026 society guidance to the 2026 CKM guideline. Stream width is proportional to weighted population; color indicates the pre-2026 drug class. The inset splits newly eligible adults by cohort. Unclassifiable adults are in the not-indicated node. Abbreviations as in Table 1.

**Table 2.** Already and newly eligible US adults under the 2026 CKM guideline, relative to society guidance. Values are weighted millions of US adults aged 30 to 79 years, with 95% CIs for newly eligible estimates. Overall-row columns are mutually exclusive. Relative increase is the newly eligible as a percentage of the already eligible. Cohort subgroups do not sum to class totals, and risk-threshold diabetes appears under both classes. 0.50 million adults (n = 53) could not be classified. T2D = type 2 diabetes; other abbreviations as in Table 1.

| Category | Eligible under either standard | Already eligible (society) | Newly eligible | Unweighted n, newly eligible | % newly among total | Relative increase, % |
| --- | --- | --- | --- | --- | --- | --- |
| GLP-1 receptor agonist | 112.58 | 105.10 | 7.48 (6.48 to 8.47) | 620 | 6.6% | 7.1% |
| Overweight, no other disease | 6.49 | 0.00 | 6.49 (5.51 to 7.47) | 472 | 100.0% |  |
| Type 2 diabetes with albuminuria | 2.27 | 2.27 | 0 | 0 | 0.0% |  |
| Type 2 diabetes at the risk threshold | 10.55 | 9.56 | 0.99 (0.75 to 1.23) | 148 | 9.4% |  |
| SGLT2 inhibitor | 44.68 | 44.68 | 0 | 0 | 0.0% | 0.0% |
| Type 2 diabetes at the risk threshold | 10.55 | 10.55 | 0 | 0 | 0.0% |  |
| CKD without diabetes | 8.10 | 8.10 | 0 | 0 | 0.0% |  |
| Overall (either class) | 119.32 | 111.84 | 7.48 (6.48 to 8.47) | 620 | 6.3% | 6.7% |

### Sensitivity Analyses

Against the full FDA label, 14.15 million adults (8.10%; 95% CI: 7.52 to 8.67; n = 1,259) were newly eligible: 6.49 million with overweight and no other cardiometabolic disease for a GLP-1RA and 7.66 million (95% CI: 6.78 to 8.54) with CKD without diabetes for an SGLT2i. Against the FDA label restricted to compelling cardiorenal disease, 25.17 million (14.41%; 95% CI: 13.64 to 15.18; n = 2,604) were newly eligible: 8.79 million (95% CI: 7.77 to 9.81) for a GLP-1RA and 18.20 million (95% CI: 16.73 to 19.67) for an SGLT2i. Counts by cohort, the corresponding clinical profiles, and the CKM stage distribution are given in Tables S5 through S8 and Figures S2 and S3. The total population the 2026 CKM guideline makes eligible for each drug class, irrespective of any prior standard, is shown in Figure 2.

**Figure 2.**
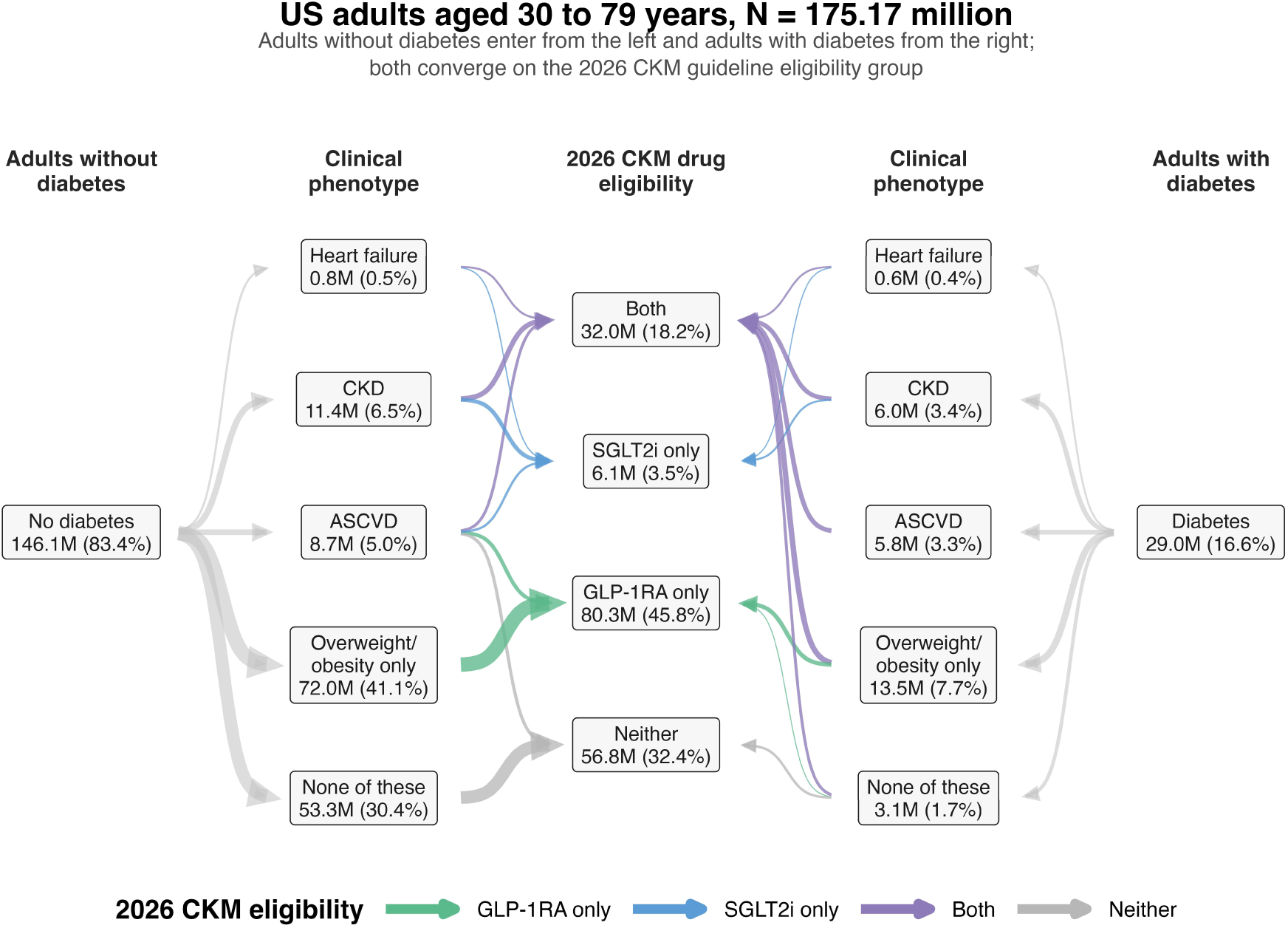
GLP-1RA and SGLT2i eligibility under the 2026 CKM guideline. Adults flow by diabetes status through mutually exclusive clinical phenotypes to total 2026 CKM guideline eligibility. Nodes show weighted millions and percentages of 175.17 million adults; connector width uses square-root scaling. Counts are totals, not increments over prior guidance (Figure 1). The SGLT2i total is lower than in Table 2 because the risk threshold excludes lower-risk adults with diabetes. Abbreviations as in Table 1.

### Clinical and Risk Profile of the Newly Eligible Population

Newly eligible adults were younger than those already eligible (mean age, 46.2 vs 52.9 years), had lower BMI (27.8 vs 33.0 kg/m²), and had lower 10-year PREVENT total-CVD risk (3.6% vs 8.1%; all P < 0.001). Among adults aged 30 to 59 years, mean 30-year risk was also lower (11.3% vs 19.6%; P < 0.001). They had less hypertension (9.0% vs 71.3%) and chronic kidney disease (0% vs 20.0%); 86.8% were at CKM stage 1, compared with 16.7% of already-eligible adults (Table 1).

Across the society, full FDA-label, and compelling-cardiorenal comparators, mean age among newly eligible adults was 46.2, 49.3, and 54.2 years, respectively; mean UACR was 6.5, 39.1, and 38.4 mg/g, and mean PREVENT total-CVD risk was 3.6%, 4.9%, and 9.7% (Table S7). Adults newly eligible for a GLP-1RA were concentrated at CKM stage 1 across all comparators, whereas those newly eligible only for an SGLT2i were concentrated at CKM stages 2 and 3; the proportion at stage 1 was 86.8%, 45.9%, and 25.8% under the three comparators, respectively (Figure S3). Profiles by drug class are given in Table S8.

### Expected Benefit of Newly Conferred Eligibility

Mean 10-year PREVENT total-CVD risk was 2.1% among newly eligible adults with overweight and no other cardiometabolic disease and 13.2% among those with type 2 diabetes at the risk threshold, against 8.1% among adults already eligible under the composite society standard. Only 6.3% (95% CI: 3.6 to 9.1) of adults with overweight and no other cardiometabolic disease reached the 10-year risk of 7.5% at which the guideline gates initiation in type 2 diabetes, compared with all of the risk-threshold diabetes cohort and 39.3% (37.8 to 40.8) of already-eligible adults. Assuming a 20% relative reduction in cardiovascular events, the 10-year number needed to treat to prevent one total cardiovascular event was 243 (95% CI: 208 to 292) in adults with overweight, 38 (35 to 41) in the risk-threshold diabetes cohort, and 138 (122 to 158) across all newly eligible adults, against 61 (60 to 63) among those already eligible (Figure 3A). Restricted to adults aged 30 to 59 years and projected over 30 years, the corresponding values were 56 (52 to 60), 13 (13 to 14), 44 (40 to 49), and 26 (25 to 26) (Figure 3B). Values for atherosclerotic cardiovascular disease and for both FDA comparators are given in Table S9.

**Figure 3.**
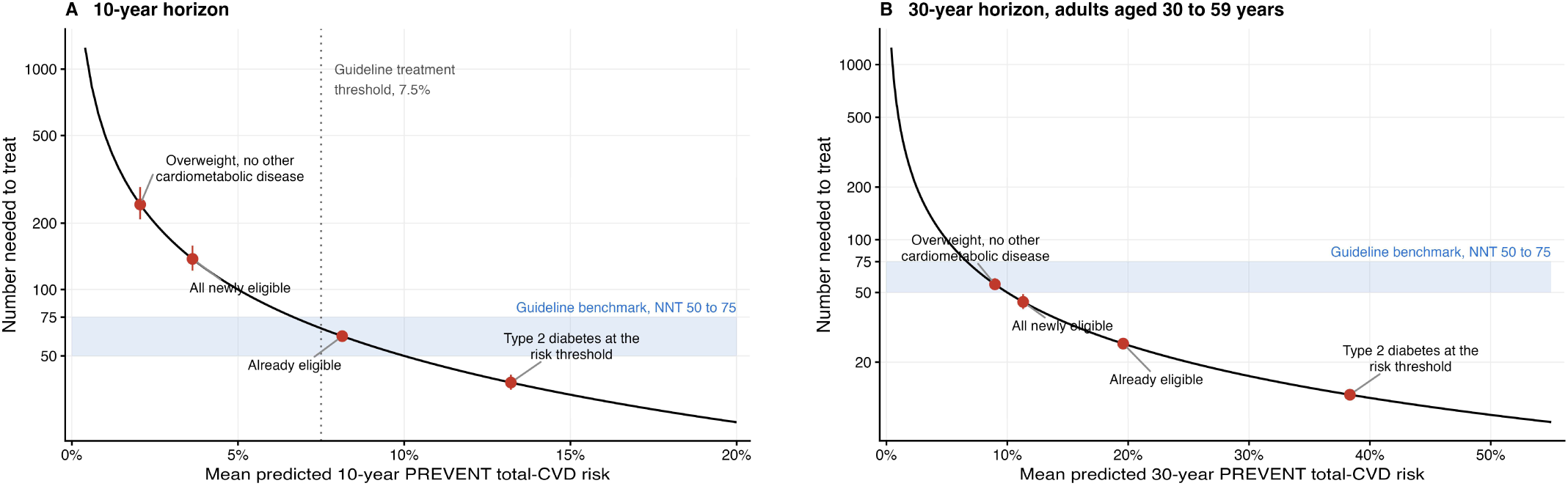
Projected number needed to treat by predicted cardiovascular risk. A, 10-year horizon. B, 30-year horizon, ages 30 to 59 years. Points show the projected number needed to treat (NNT; 95% CI) against mean PREVENT total-CVD risk; the curve assumes a 20% relative risk reduction. Shading marks the guideline NNT benchmark of 50 to 75; the dotted line marks the 7.5% diabetes threshold. Values are in Table S9.

## DISCUSSION

In this nationally representative study, the 2026 CKM guideline made 7.48 million US adults newly eligible for a GLP-1RA relative to pre-2026 society guidance and none newly eligible for an SGLT2i (Figure 4). These newly eligible adults were predominantly at CKM stage 1 and had lower predicted cardiovascular risk than those already eligible. Estimates varied substantially by comparator, ranging from 7.48 million against the society composite to 25.17 million against FDA labeling restricted to compelling cardiorenal disease, largely reflecting differences in how type 2 diabetes was treated across reference standards.

**Figure 4.**
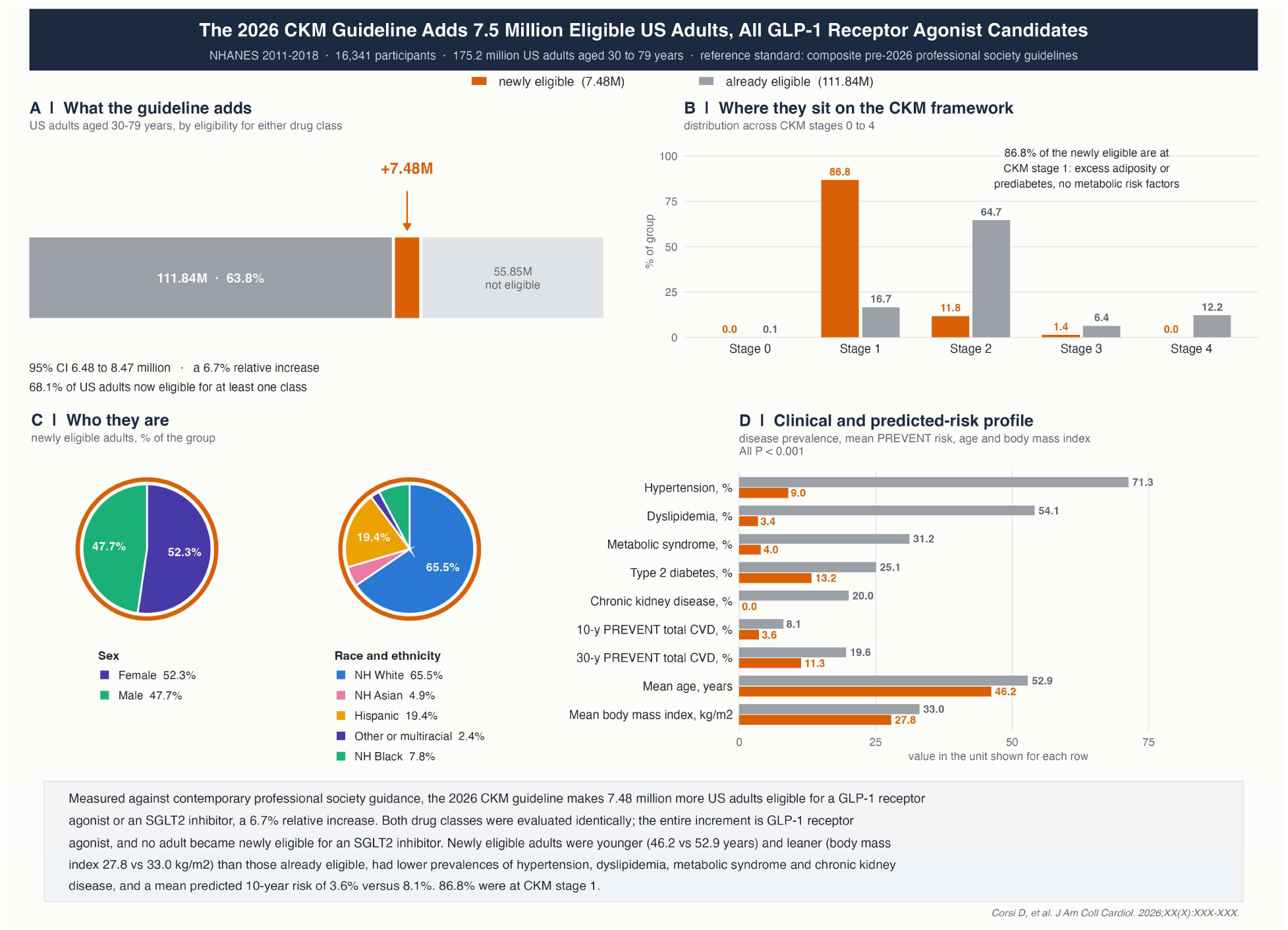
2026 CKM Guideline Adds 7.5 Million Adults, All GLP-1RA Candidates. Survey-weighted NHANES 2011 to 2018 estimates for adults aged 30 to 79 years, relative to pre-2026 society guidance. A, Eligibility before and after the 2026 CKM guideline. B, CKM stage distribution. C, Sex and race and ethnicity of newly eligible adults; Hispanic combines Mexican American and other Hispanic. D, Disease prevalence, mean PREVENT risk, age, and BMI. Thirty-year risk is restricted to ages 30 to 59 years.

This variation reflects differences in comparator definitions. Two definitional choices account for it: whether type 2 diabetes alone is an indication, which explains the entire difference between the compelling-cardiorenal and full-FDA comparators (25.17 vs 14.15 million), and how CKD is defined, which explains most of the further narrowing to the society composite (14.15 to 7.48 million), because KDIGO criteria already capture every adult meeting the guideline’s albuminuria threshold. Label permission is the least informative reading: FDA labeling has long allowed both classes in any adult with type 2 diabetes, whereas the guideline directs that PREVENT risk be estimated and treatment offered for cardiovascular, kidney, and mortality benefit rather than glycemic control alone. This shift toward earlier organ-protective therapy is consistent with the 2026 ADA/European Association for the Study of Diabetes (EASD) consensus report, which advocates earlier initiation of GLP-1-based therapies and/or SGLT2 inhibitors in type 2 diabetes, potentially at diagnosis, without requiring a specific predicted cardiovascular risk threshold.^22^ Even against the most conservative comparator, which counted adults as already eligible if they met any of the three standards, the increment consisted entirely of GLP-1RA therapy for overweight at CKM stage 1, so its concentration in lower-risk adults is robust. Prior coverage also explains why the writing committee’s finding that 83% of US adults with diabetes at CKM stages 2 to 3 were eligible for cardioprotective therapy^21^ does not describe a largely new population: only 0.99 million such adults were newly eligible against the society composite, because prevalent cardiorenal disease and ADA high-risk criteria already captured the rest. An analysis of the 2026 dyslipidemia guideline similarly found that newly statin-eligible adults were younger and at lower risk.^6^

The projected benefit is dependent on the timeline for which it is measured. The guideline gates initiation in adults with type 2 diabetes at a 10-year PREVENT-CVD risk of 7.5% or greater but sets no risk threshold for its CKM stage 1 obesity recommendation; applied to newly eligible adults with overweight, that threshold would select only 6.3% (95% CI: 3.6 to 9.1), none of which would have met SELECT entry criteria.^23^ Assuming the 20% relative reduction in major adverse cardiovascular events used in the guideline’s own benefit calculations,^21^ consistent with SELECT participants with a BMI below 30 kg/m²^24^ and a meta-analysis of 12 trials,^25^ the 10-year number needed to treat in this group is about 4 times that among adults already eligible and exceeds the range of 50 to 75 below which guidelines have regarded preventive therapy as justified;^21^ over 30 years, it falls within that range. Longitudinal data suggest that many adults with metabolically healthy overweight develop metabolic disease and higher risk,^3,26^ and this process may begin before conventional risk factors appear: high-sensitivity C-reactive protein is nearly twice as high at CKM stage 1 as at stage 0,^27^ so the label metabolically healthy may understate risk in this group. In addition, the data on metabolically healthy adults do not account for risk of developing heart failure with preserved ejection fraction (HFpEF).^28^ Whether treating now, rather than at metabolic transition or unappreciated HFpEF risk, delivers the projected benefit is untested: no completed outcome trial has enrolled these adults, SURMOUNT-MMO requires the risk factors this cohort lacks,^29^ and trials in this population are needed. Value is similarly uncertain. The guideline judged GLP-1-based therapy at CKM stage 1 not cost-effective at 2025 US prices, and a microsimulation in a heavier, higher-risk, label-eligible US population estimated incremental cost-effectiveness ratios of $467,676 (semaglutide) and $197,023 (tirzepatide) per quality-adjusted life-year versus lifestyle intervention,^30^ suggesting uncertain value in this lower-risk group. Because these adults fall outside FDA labeling for chronic weight management, coverage is unlikely and out-of-pocket costs could widen disparities in access,^31^ so structured lifestyle intervention should remain foundational.

Implementation remains a major challenge: an estimated 105.10 million US adults were already eligible for a GLP-1RA and 44.68 million for an SGLT2i under the society composite, far outnumbering the increment. Among US adults with diabetes, GLP-1RA and SGLT2i use has been lower with CKD,^32^ while approximately half of adults at CKM stage 2 or higher with hypertension or hyperlipidemia were untreated.^33^ The composition of the increment also warrants interpretation. Non-Hispanic Black adults comprised 7.8% of the newly eligible but 12.9% of the already eligible, whereas Hispanic adults were modestly overrepresented (19.4% vs 15.4%; P < 0.001 for the overall distribution). This reflects prior inclusion rather than new exclusion: the new recommendation applies only to the 27.0 to 29.9 kg/m² band, and adults with a BMI of 30 kg/m² or above, among whom non-Hispanic Black women are overrepresented,^34^ were already eligible. Equitable benefit will therefore require attention to implementation among both previously and newly eligible adults, monitored by race and ethnicity, sex, social determinants of health, and kidney disease status.

## LIMITATIONS

First, disease constructs relied on self-report or single measurements. Under-ascertainment of self-reported ASCVD and heart failure overstates the newly eligible population by understating the already eligible, and a single urine albumin-to-creatinine ratio cannot establish three-month persistence and likely overstates CKD, with effects that differ by comparator. Cardiac biomarkers, cardiac imaging, atrial fibrillation, and peripheral artery disease were unavailable, so subclinical cardiovascular disease could not be operationalized and stages 3 and 4 may be underestimated; stage 4 could not be split into 4a and 4b because kidney replacement therapy was not ascertained. Second, the GLP-1RA recommendation for heart failure with preserved ejection fraction could not be estimated because NHANES does not measure ejection fraction, so the estimates exclude any adults that recommendation would add. Third, diabetes could not be subtyped, so adults with type 1 diabetes are included in the diabetes-defined cohorts and comparators; because type 1 diabetes is a small minority at these ages, misclassification is likely modest. Fourth, obstructive sleep apnea and metabolic dysfunction-associated steatohepatitis could not be ascertained, so some adults counted as newly eligible may already have qualified. Fifth, PREVENT could not be computed for 1,711 of 16,341 participants (10.5%) because of missing laboratory values, so the risk-threshold cohort is conditioned on computability (Supplemental Methods). Sixth, eligibility was classified without regard to current pharmacotherapy, so some adults counted as newly eligible may already be receiving one of these agents, and the estimates describe the population a guideline identifies rather than the population whose treatment would change. Seventh, the assumed relative risk reduction has not been established in adults without cardiovascular disease: in LEADER, liraglutide showed no apparent benefit among participants with cardiovascular risk factors alone (hazard ratio, 1.08; 95% CI: 0.84 to 1.38),^35^ and in SELECT, changes in body weight, blood pressure, cholesterol, and glycemia jointly explained only about one-third of the benefit,^36^ so the mechanism, and its applicability to adults without established atherosclerosis, remains uncertain. Absolute benefit in adults with overweight and no other cardiometabolic disease is therefore less certain than its predicted risk.

## CONCLUSIONS

Relative to contemporary professional society guidance, the 2026 CKM guideline newly identified 7.48 million US adults aged 30 to 79 years (95% CI: 6.48 to 8.47) as candidates for pharmacotherapy, a 6.7% increase over the 111.84 million already eligible. All became eligible for a GLP-1 receptor agonist; most had overweight without other cardiometabolic disease. None became newly eligible for an SGLT2 inhibitor. Expected benefit in this group is modest over 10 years and more favorable over 30. Because these adults were generally not represented in phase 3 obesity pharmacotherapy trials, additional outcome and cost-effectiveness data are needed to better define the potential benefits and inform treatment decisions in this population.

## Data Availability

The datasets analyzed during the current study are publicly available in the National Health and Nutrition Examination Survey (NHANES) repository maintained by the Centers for Disease Control and Prevention (CDC) / National Center for Health Statistics (NCHS).

https://www.cdc.gov/nchs/nhanes/index.htm

## ACKNOWLEDGMENTS

Claude (Anthropic) was used to assist with manuscript editing and analysis-code review. The authors reviewed all output and take full responsibility for the content of this article.

## SUPPLEMENTAL MATERIAL

Supplemental Methods

Tables S1–S9

Figures S1–S3

## NON-STANDARD ABBREVIATIONS AND ACRONYMS

ASCVD: atherosclerotic cardiovascular disease
CKD: chronic kidney disease
CKM: cardiovascular-kidney-metabolic
eGFR: estimated glomerular filtration rate
GLP-1RA: glucagon-like peptide-1 receptor agonist
NHANES: National Health and Nutrition Examination Survey
PREVENT: Predicting Risk of Cardiovascular Disease EVENTs
SGLT2i: sodium-glucose cotransporter 2 inhibitor
T2D: type 2 diabetes
UACR: urine albumin-to-creatinine ratio

